# Study protocol for the Multimodal Approach to Preventing Suicide in Schools (MAPSS) project: A regionally based feasibility trial of an integrated response to suicide risk among UK secondary school pupils

**DOI:** 10.1101/2024.04.17.24305955

**Authors:** Emma Ashworth, Molly McCarthy, Sio Wynne, Jo Robinson, Samuel McKay, Steven Lane, Gerry Richardson, Neil Boardman, Kate Henderson, Vivienne Crosbie, Neil Humphrey, Sian York, Maria Michail, Damian Hart, David Clacy, Mani Jalota, Pooja Saini

## Abstract

**Background:** Suicide is the leading cause of death of children and young people under 35 in the UK, and suicide rates are rising in this age group. Schools are considered an appropriate and logical setting for youth suicide prevention activities, with universal, selective, and indicated approaches all demonstrating efficacy. Given that international best practice recommends suicide prevention programmes combine these approaches, and that to date this has not been done in school settings in the UK, this study aims to evaluate the feasibility of delivering a suicide prevention programme incorporating universal, selective, and indicated components in UK schools.

**Methods:** This study is a feasibility cluster-randomised controlled trial (RCT) of an adapted version of the Multimodal Approach to Preventing Suicide in Schools (MAPSS) programme. The programme, initially developed in Australia, involves delivering universal psychoeducation to all pupils, screening them for suicide risk, and delivering Internet-based Cognitive Behavioural Therapy (Reframe IT-UK) to those students identified as being at high-risk for suicide. The programme will be trialled in six secondary schools in Northwest England and will target Year 10 students (14- and 15-year-olds). The primary aims are to assess: 1) the acceptability and safety of delivering MAPSS in a school setting in the UK; 2) the social validity of the MAPSS programme; and 3) the feasibility of delivering a large-scale, appropriately powered, cluster-RCT and economic evaluation of this intervention in the future. Secondary aims are to assess changes over time in mental health and wellbeing outcomes.

**Discussion:** This study is the first to evaluate a suicide prevention programme comprising universal, selective, and indicated components in UK schools. If the programme is found to be feasible, it could be more widely tested in schools and may ultimately lead to reduced rates of suicide and suicidal behaviour in young people.

## Introduction

Rates of suicidal crisis among children and young people (CYP) are on the rise, with suicide rates per 100,000 adolescents having increased by 7-9% per year since 2010 (1), and suicide now being the leading cause of death of young people under 35 (2). In Northwest England, there has been an increase in attendances to Emergency Departments for CYP in suicidal crisis and/or for self-harm (3), and the number of CYP presenting is significantly worse than the UK average (4). Suicidal ideation and behaviour are associated with a host of negative outcomes including risk of future suicide (5). The impact of suicide on a young person’s family, friends, and wider community can also be devastating, and increases their own risk of suicide (6). There is, therefore, an urgent need to develop and test acceptable and effective approaches to preventing suicide in this population.

Schools are an appropriate setting for the delivery of mental health prevention programmes, offering a ‘universal access point’ to many CYP, and have been identified as important locations for suicide prevention and early intervention activities. Although school wellbeing staff may be able to provide support to pupils, CYP are often reluctant to seek help from professionals, preferring informal sources of support (7). School-based prevention efforts must therefore not only target school staff, but also fellow pupils. Historically, there has been a reluctance to deliver suicide prevention efforts to pupils, due to concerns about potentially iatrogenic impacts. However, increasing evidence suggests that it is safe to do (8,9). According to international best practice, suicide prevention programmes should incorporate universal, selective, and indicated approaches. Such approaches have shown promise in both community and school settings (10) but, to date, only one study (11) is applying rigorous methodology to evaluate short- and longer-term cost-effectiveness of an intervention comprising universal, selective, and indicated elements in schools.

The Multimodal Approach to Preventing Suicide in Schools (MAPSS) project, a suicide prevention intervention in Australia, has demonstrated feasibility and acceptability and is currently undergoing a randomised controlled trial (RCT) in Melbourne (11). The MAPPS intervention consists of three parts: suicide prevention lesson for all pupils, risk screening, and online cognitive behavioural therapy (CBT; ‘Reframe-IT’) for those deemed to be at high risk for suicide ideation. Further details for each element are provided below. Training is also provided for school staff and parents. Suicide prevention lessons, such as those included in MAPSS, have been evaluated in youth populations (8,9), and ‘Reframe-IT’ has also been found to be associated with reduced suicidal ideation, depression, and hopelessness in Australian CYP (8,12). However, cultural transferability of interventions cannot be assumed (13); interventions that have worked in one setting or context too often do not work across other settings, particularly in school contexts (14), given the wide ranging contextual and cultural factors influencing implementation (15). Further to this, if an intervention does not have high social validity, meaning that it is not viewed as acceptable, useful, and feasible by intervention deliverers (e.g., school staff) and/or recipients (e.g., pupils), then it is likely to fail (16,17). Therefore, before any suicide prevention interventions are delivered at-scale in UK schools, the social validity of such interventions should be established, along with any necessary cultural or contextual adaptations, to ensure success.

A recent scoping study of MAPSS for UK schools (18,19) interviewed CYP, school staff, parents, and health professionals. All participants advocated the importance of school-based suicide prevention and gave feedback on the changes needed to adapt the MAPSS intervention for the UK. An adapted version of MAPSS has since been co-developed with CYP and professionals, and examined in a pilot study in two schools. Findings showed it was feasible to recruit schools, have suicide prevention training delivered to school staff and suicide awareness sessions to parents, for researchers to conduct surveys within the school setting at study timepoints, to identify pupils who may be at risk of suicide, to deliver suicide prevention lessons with Year 10 pupils, and to recruit pupils to test the online CBT therapy programme, Reframe IT-UK.

### Aims and Hypotheses

The proposed study aims to build on the aforementioned work, employing a feasibility RCT design of an adapted version of MAPSS across six schools in Northwest England to assess: 1) the acceptability and safety of delivering MAPSS in a school setting in England; 2) the social validity (feasibility, utility, and acceptability) of the MAPSS intervention; and 3) the feasibility of delivering a large-scale, appropriately powered, cluster-RCT and economic evaluation of this intervention in the future. Secondary aims are to assess changes over time in mental health and wellbeing outcomes for pupils taking part in MAPSS. As this is a feasibility study, no formal hypothesis testing will be undertaken.

## Materials and Methods

### Study Design

The study is led by researchers at Liverpool John Moores University (LJMU), UK, and will involve schools across Cheshire and Merseyside in Northwest England. This feasibility study is funded by the National Institute for Health and Care Research (NIHR) Public Health Research Programme (NIHR156862), and delivery of the MAPSS interventions is commissioned by Cheshire and Merseyside Public Health Collaborative (CHAMPS). It will be conducted over a two-year period (Feb 2024-Feb 2026), with three months for set up, 18 months feasibility, and three months consolidation. This study adopts a multiphase feasibility cluster-RCT design (ISRCTN: 41891301; see SPIRIT and CONSORT diagrams, Figures 1 and 2), consisting of a treatment as usual (TAU) arm and an intervention (MAPSS) arm. Schools allocated to the TAU arm will be offered MAPSS at the end of the trial, if it is considered safe and acceptable for use.

**Figure 1.**
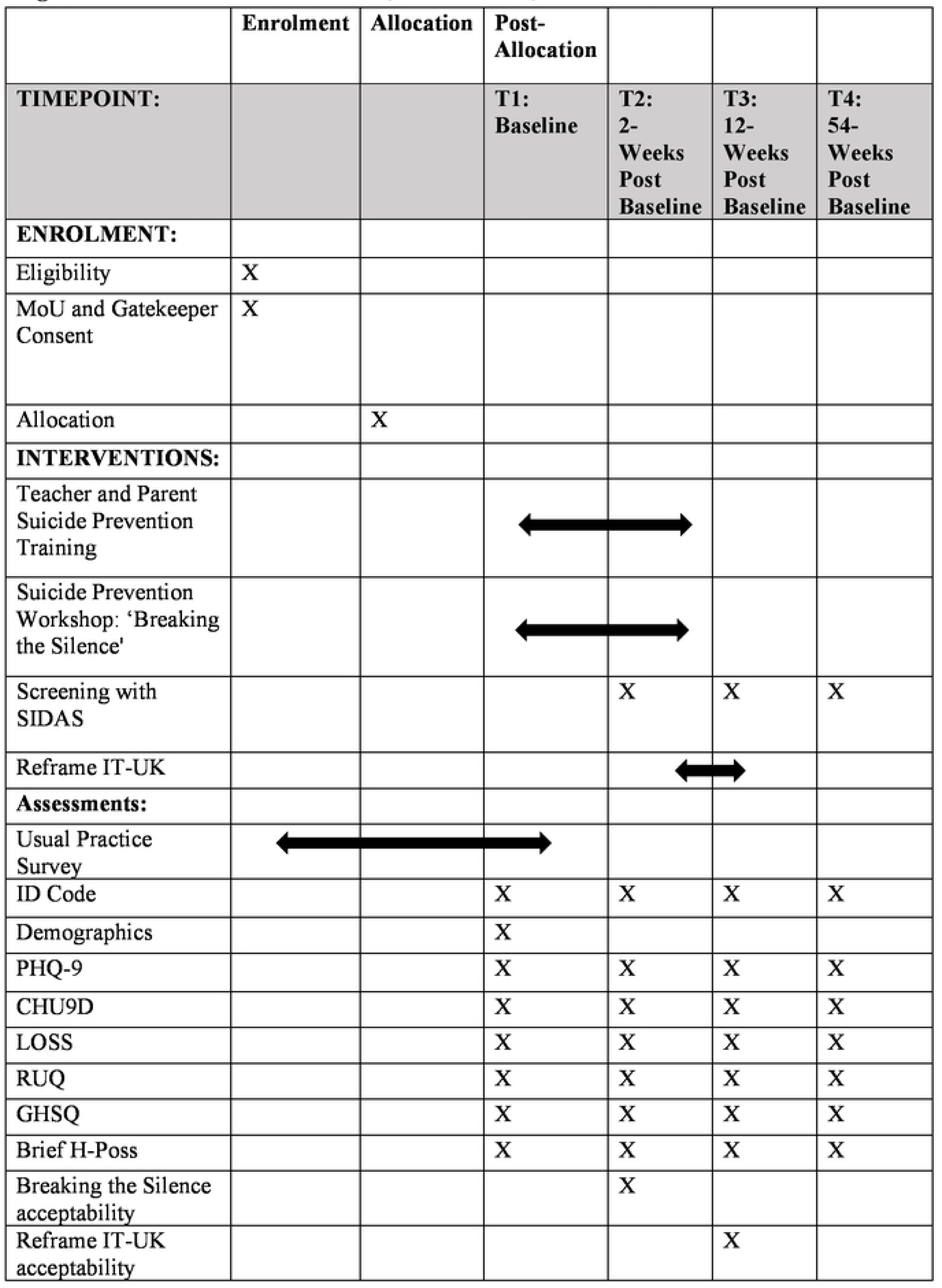
SPIRIT schedule of enrollment, interventions, and assessments.

**Figure 2.**
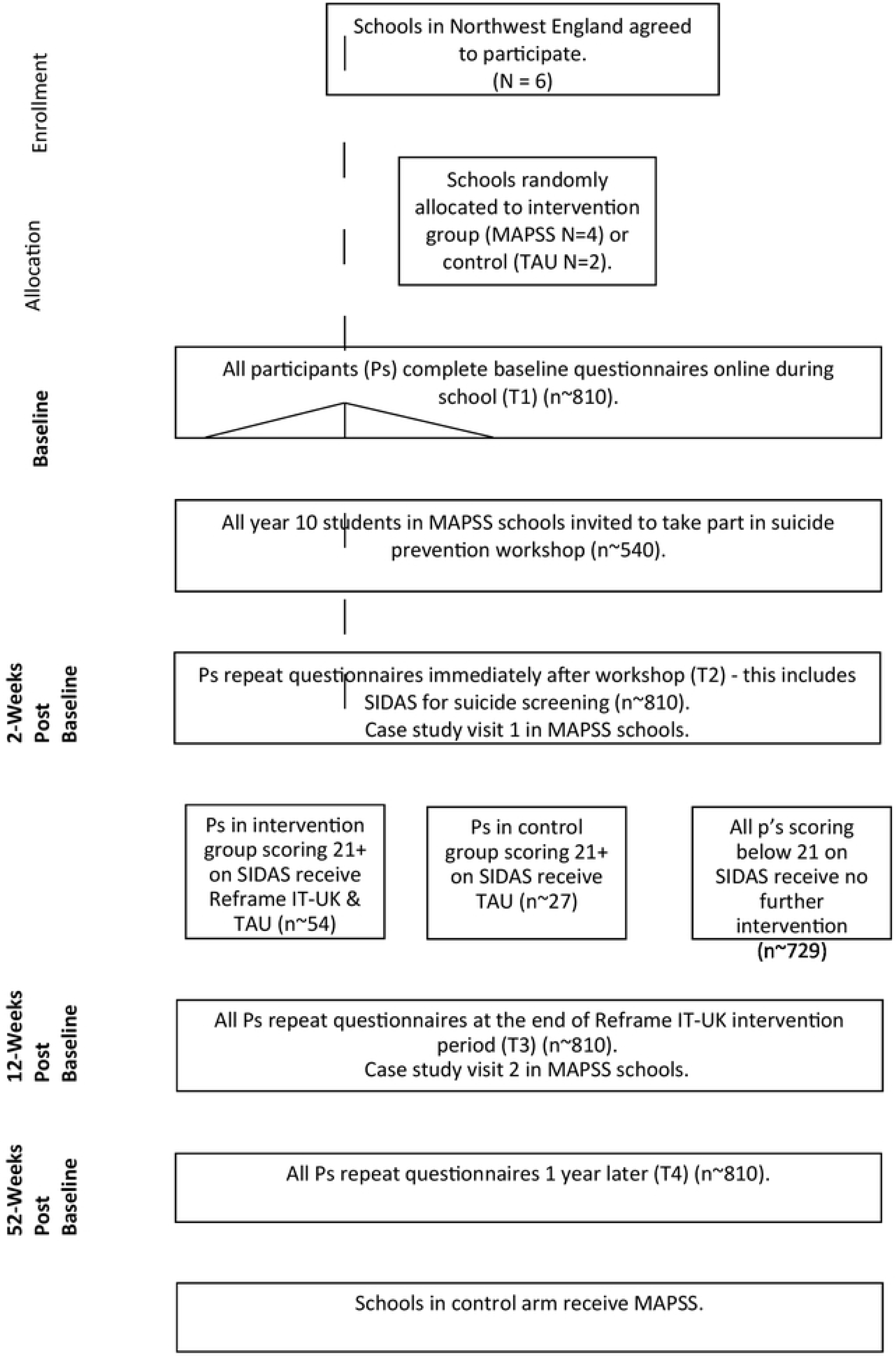
MAPSS anticipated CONSORT diagram.

The study consists of three work packages (WP):

**WP1:** A three-month set-up stage to: i) make any adjustments to the interventions as determined from the pilot study, ii) recruit staff, and iii) spend dedicated time recruiting schools from a diverse range of backgrounds, with three schools recruited in the first three months and the remaining three schools within nine months, to enable alignment with school academic terms.

**WP2:** An 18-month feasibility cluster-RCT evaluating MAPSS in six schools. Four schools will be randomised to receive MAPSS (intervention arm), and two schools will be randomised to continue with usual practice (TAU control arm). Surveys will be completed by both arms at all timepoints. This will include i) baseline surveys, ii) universal suicide prevention lesson (‘Breaking the Silence’ – see further details below) in the intervention arm, iii) survey and screening two weeks after the suicide prevention lesson, iv) an indicated Reframe IT-UK CBT programme in the intervention arm for pupils identified as high suicide risk, and usual care in control arm, v) surveys two-weeks after Reframe IT-UK, and vi) follow-up surveys 12-months post-baseline.

**WP3.** A parallel process evaluation, to establish perceptions of social validity of the programme for use in the UK, and the appropriateness of the research design for effectiveness trials.

### Participants and Recruitment

Participant recruitment will begin in February 2024. Expressions of interest will be sought from schools across Cheshire and Merseyside via Local Authority contacts. To ensure recruitment of a diverse range of schools and CYP, purposeful maximum variation sampling will be used if more than six schools express an interest in taking part. This is widely used in research to pragmatically identify and select participants that are effective in addressing the research aims, while also maximising diversity and limiting bias (20). The key characteristics we will seek variation on include: rural/urban status, proportion of ethnic minority pupils, schools’ deprivation levels (IDACI), schools’ academic achievement (proportion of pupils achieving benchmarks GCSE grades).

Selected schools will be provided with a gatekeeper information sheet and opt-in consent form to sign and return to the research team, along with a memorandum of understanding (MoU). Schools will be randomised once these documents are received. Information sheet and opt-out consent forms will then be sent to parents/carers of pupils in the participating schools. Finally, assent will be sought from young people during baseline survey completion. The sample will consist of approximately 810 adolescent pupils in Year 10 (aged 14-15), recruited from six mainstream secondary schools across Cheshire and Merseyside. The six schools will be randomly assigned to one of two arms as part of the cluster-RCT: intervention (n=4) or control (TAU) arm (n=2). Year 10 pupils in the intervention schools will receive: 1) a suicide prevention psychoeducation lesson (n∼540 pupils); and 2) pupils scoring 21 or above on the Suicide Ideation Attributes Scale (SIDAS)^33^ or indicating past suicide ideation will also be offered Reframe IT-UK plus TAU (n∼54 pupils). Those in the control arm will receive TAU only (n∼27 pupils).

As this is a feasibility study, a sample size calculation is not needed. In line with similar work (21), we will recruit six schools to provide sufficient variation of schools and pupil numbers in which to test recruitment, retention, acceptability of the intervention, and feasibility of the research design for evaluation. This is based on the assumption of a potential attrition rate of 50% at T3 (the primary outcome point for Reframe IT-UK). This is common for feasibility trials, realistic in terms of recruitment, and allows adequate precision in estimating rates (e.g., attrition, adverse events) relevant to trial outcomes (21). This would allow an overall attrition rate of 50% to be estimated with 95% confidence intervals of +/- 12%, or 16% for a single arm. This sample size is also adequate for estimating relevant analysis parameters such as the standard deviation of effects, which are needed for determining the feasibility of a later efficacy trial. Based on findings from the pilot study, we anticipate that 10% of pupils will score in the at-risk range in the screening and will thus be eligible for Reframe IT-UK. Recruitment rates will be monitored by collecting data on:

1. The proportion of eligible young people who consented,
2. The number of participants recruited during the recruitment stage of feasibility compared with the target,
3. Assessment of contamination of MAPSS programme in control schools,
4. Assessment of CYP satisfaction with intervention and outcome measures.

This will provide evidence on recruiting to trials in school settings, as well as informing the full trial design.

### Inclusion Criteria

Pupils are eligible to take part if:

1. They are in Year 10 at school, aged 14+ years,
2. They attend a mainstream secondary school or pupil referral unit in Cheshire or Merseyside,
3. Their parents do not withdraw consent,
4. They provide assent,
5. Their school is willing to deliver MAPSS (if allocated), facilitate data collection, and completes a gatekeeper consent form and MoU,
6. For Reframe IT-UK only: they score 21 or above on the SIDAS or indicate thoughts of suicide in the past month at T2 data collection.

### Randomisation

The unit of randomisation is the schools. Six schools will be randomised to one of two study groups after they have returned the appropriate documentation to secure their place in the trial. We will be comparing both arms to test which, if any, is better. To create balance in terms of deprivation, ethnicity, rurality and educational outcomes, a minimisation algorithm will be used at an intervention-to-control ratio of 2:1 across Cheshire and Merseyside. Two schools will be allocated to the control arm and four schools will be allocated to the intervention arm by a university statistician, who is independent of the study and blind to school identities (blinding of schools and participants themselves is not possible due to obvious differences in intervention delivery). Methods of allocation concealment and randomisation processes will follow CONSORT (22). Schools will be randomised to receive a suicide prevention lesson and Reframe IT-UK and TAU, or TAU only, via a random sequence generation computer algorithm. Researchers completing study assessments will be masked to intervention allocation. The trial will follow an Intent-To-Treat (ITT) protocol.

### Withdrawal Criteria

Schools or pupils can withdraw from the trial at any time. If an individual pupil withdraws, no further action will be taken and their data will be deleted. If a school withdraws prior to intervention delivery beginning, we will seek to replace the school. If a school withdraws after this point, we will not seek to replace the school and the trial will continue. We will aim to complete exit interviews with any schools that withdraw, to ascertain their reasons for withdrawal. The trial may be prematurely stopped if a significant adverse event occurs as a result of the trial procedures.

### Procedure

Following recruitment, all participants will complete a suite of quantitative measures online in school at four time-points: baseline (T1); 2-4 weeks post-baseline (after suicide prevention lesson; T2); 12 weeks post-baseline (after Reframe IT-UK; T3); and 1-year post-baseline (T4). Schools will be asked to provide IT access for pupils to complete the surveys and will be provided with a detailed support pack for completing the measures with pupils, including links to the surveys, age-appropriate lesson plans, PowerPoint slides, and glossary. A member of the research team will visit the school at T1 to explain the study to the participants and ensure they are fully informed about the study and their rights before they assent. Surveys will not be anonymous as pupils will need to be monitored for risk and screened for potential participation in Reframe IT-UK.

At T1, a minimum of six staff from each school (intervention and control) will receive training from Papyrus Prevention of Young Suicide (a national charity), known as ‘Suicide Prevention – Overview Tutorial’ (SP-OT), to ensure staff are equipped to manage any risk identified from the screening. SP-OT is delivered in a single session over 1.5 hours online. Papyrus will also provide an online information session, ‘Suicide Prevention - Awareness, Resource, Knowledge’ (SP-ARK), along with support packs, for parents of all children in Year 10 at each participating school. #MyGPGuide; a guide for CYP with lived experience of self-harm and suicidality will be shared with schools and families (23).

Between T1 and T2 data collection, intervention schools will receive the suicide prevention lesson. At T2, pupils will complete the SIDAS and a single-item question relating to suicide ideation in the past month. The survey system will flag participants who score in the at-risk range for suicidal ideation and the research team will then contact the school about these pupils to determine eligibility for participation in the Reframe IT-UK in intervention schools. The details of at-risk pupils will be passed on to schools the same day as survey completion, and they will be asked to follow up with these pupils (within 24 hours for high-risk and five working days for medium-risk) and adhere to their usual safeguarding procedures. At-risk pupils will then be offered the intervention and TAU in intervention schools, or TAU only in control schools. Pupils receiving the Reframe IT-UK intervention will complete the eight modules in the 10 weeks between T2 and T3 (i.e., approximately one module per week, while allowing an additional two additional weeks to account for disruptions such as school holidays). Completion of Reframe IT-UK takes place during school time, and a pastoral member of staff will sit with and support the young people while they complete the programme and provide any immediate support if they report suicide risk either directly to the staff member or via the Reframe IT-UK mood check-ins.

### Intervention

1. A **suicide alertness training workshop, ‘**Breaking the Silence’, developed for and tested with young people, will be delivered. This psychoeducation lesson comprises a single 3-hour face-to-face workshop, designed to help participants understand suicide warning signs in themselves and others, gain knowledge about sources of support, and signpost others. Suicide prevention lessons will be delivered by trained facilitators from Grassroots Suicide Prevention (a national charity) to classroom-sized groups of pupils (maximum 30 pupils per session with at least one teacher present). A researcher will also attend where possible, to complete field notes.
2. The **screening** will take the form of self-report measures embedded into the questionnaires at each timepoint (excluding baseline). Researchers will inform schools after each timepoint of any pupils who are assessed to be at risk. Pupils who report suicidal ideation within the past four weeks (SIDAS score of 21 or higher) or any level of current suicidal ideation (single multiple-choice item) will be flagged by the research team and followed up by the school safeguarding lead.
3. **Reframe IT-UK** has been adapted from the Reframe-IT intervention developed in Australia (8,12). It comprises eight 20-minute online self-guided CBT modules. It follows the stories of two young people who make video diaries about their day-to-day life and their experiences of feeling suicidal. An adult “host” character guides the user through the module and activities. Each module contains two “activities” based on standard CBT exercises. Users progress sequentially through the content, with modules automatically unlocking when the preceding module is complete. There is also a message board through which the participant can communicate with a moderator, a mood diary function, and a series of factsheets and information on local and national helplines and services. Adaptations from the Australian version included surface-level changes, such as using British actors for the video diaries, modernising content (e.g., discussions of the COVID-19 pandemic), providing UK sources of support, and making completion of the safety plan compulsory prior to the first module unlocking.

Participants in schools randomised to the control group will receive TAU (e.g., from the school nurse or external mental health services), based on the typical provision at each school. The pastoral staff will be asked to record what TAU comprises in each school through the completion of a ‘usual practice’ survey prior to randomisation and at T4, to establish programme differentiation, any changes over time, and to control for any compensatory rivalry that may occur over the course of the trial. To ensure safety and appropriate support in the event of pupils being flagged as at-risk in the control group, schools who do not engage with the SP-OT teacher training will be unable to progress through the trial.

### Assessment of Outcomes

Primary outcome measures for CYP are listed in Table 1.

**Table 1.**
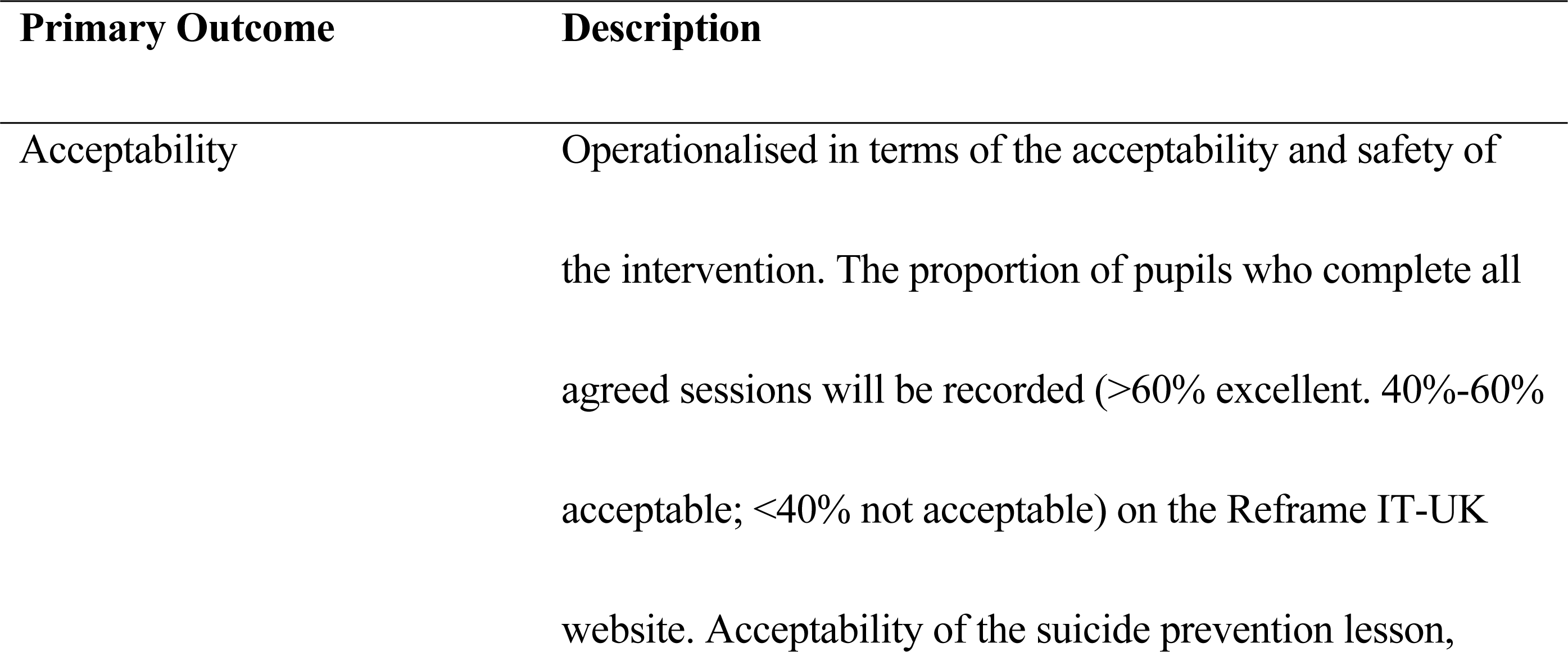
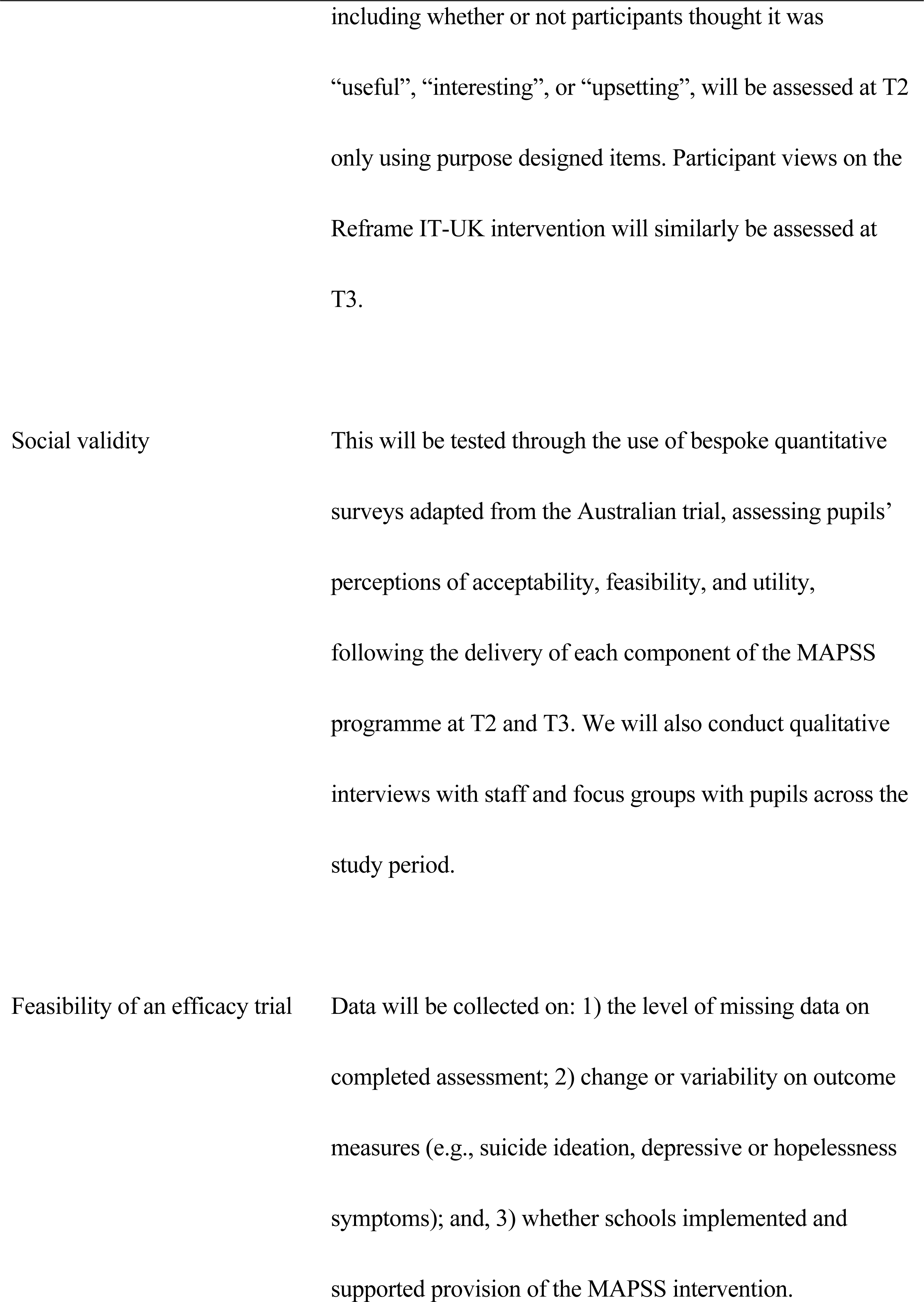
Description of primary outcome measures Primary Outcome Description.

Secondary outcome measures include:

1. Change in **past four-week suicidal ideation** at T3 and T4, compared to T2 and T1, will be assessed via the SIDAS (24). The SIDAS is a self-report measure designed to screen individuals in the community for presence of suicidal thoughts and assess the severity of these thoughts. It comprises five items, each targeting an attribute of suicidal thoughts: frequency, controllability, closeness to attempt, level of distress associated with the thoughts, and impact on daily functioning. Responses are measured on a 10-point scale. Total SIDAS scores are calculated as the sum of the five items, with controllability reverse scored, with total scale scores ranging from 0 to 50. Items are coded so that a higher total score reflects more severe suicidal thoughts. Scores over 21 (the clinically significant threshold for suicide ideation) will determine eligibility for Reframe IT-UK (Or TAU).
2. Change in **symptoms of depression** at T3 and T4, compared to T2 and T1, will be assessed using the Patient Health Questionnaire – 9-item (PHQ-9) version (25). Participants will be asked to indicate how often they have been bothered by nine problems over the past two weeks. Each item is rated on a four-point Likert scale ranging from 0 (“not at all”) to 3 (“nearly every day”). Scores are summed such that the potential range is 0–27, with higher scores indicative of greater distress. The final question of the PHQ-9 asks participants to indicate the number of days they have had thoughts that they would be better off dead, or hurting themselves in some way. If they indicate any suicidal thoughts, participants will be presented with a purpose-designed item, asking them to indicate which of the following describes the level of suicidal ideation they are experiencing: “Mild suicidal thoughts with no plan or intent to act”; “Moderate suicidal thoughts with a rough plan and some intent”; “Severe suicidal thoughts with a specific plan and intent to act”. Responses on this item will be used to determine eligibility for Reframe IT-UK (Or TAU).
3. Changes in **hopelessness** at T3 and T4, compared to T2 and T1, will be assessed using the Brief-H-Pos, a two-item positively worded measure of hopelessness (26). Respondents indicate agreement on a five-point scale (range 2–10), with higher scores indicating higher hopelessness.
4. Differences in **health service use** and other resource use (education and local authority) will be assessed at T1, T2, T3, and T4, using the Resource Use Questionnaire (RUQ), a bespoke questionnaire adapted from the Young Mind Matters Service Use questionnaire (27). The RUQ assesses use of mental health professionals and services, mental health-related hospitalisations, medication use for mental health reasons, and school-based mental health services. These data will be used for the economic evaluation.
5. Changes in **intentions to seek help** at T3 and T4, compared to T2 and T1, will be assessed using part two of the General Help Seeking Questionnaire (GHSQ; 26). The GHSQ presents participants with a list of potential sources of help and asks them to indicate the likelihood that they would approach that source if they were experiencing suicidal thoughts on a five-point scale (very unlikely-very likely). Higher scores indicate greater levels of intended help-seeking.
6. Change in **health-related quality of life** during the trial (T1, T2, T3, T4) will be assessed using the Child Health Utility–9 (CHU9D; 27). The CHU9D can be used to derive quality-adjusted life years (QALYs). These data will be used in combination with data from the health service use measure to assess the feasibility of the full economic evaluation. The CHU9D is a multi-attribute utility instrument suitable for young people aged seven–17 years. It comprises a short questionnaire alongside a set of preference weights using general population values. The questionnaire has nine items with five response levels per item.
7. Change in **suicide literacy** at T2, T3 and T4, compared to T1, will be assessed using an adapted version of the Literacy of Suicide Scale (LOSS) – short form (30). This contains 12 statements rated on a “true/false/don’t know” scale. The correct response for items 1, 3, 4, 5, 7, 8, and 10 is “false” while items 2, 6, 9, 11, and 12 are correctly answered “true”. The scale provides a total literacy score (percent correct), where higher scores indicate greater suicide literacy.
8. School staff (key contact or safeguarding lead) will complete a purpose-designed **usual practice** survey at T1 and T4, to ascertain current provision (i.e., establish a clear counterfactual), identify the level of programme differentiation, and to account for any potential compensatory rivalry or contamination in control schools.

### Implementation and Process Evaluation

A parallel qualitative implementation and process evaluation (IPE) will be conducted alongside the RCT. Longitudinal case studies will be conducted of the four schools randomised to receive MAPSS. The case studies will explore inter-related issues of 1) social validity of MAPSS and 2) *how* MAPSS was implemented and *why* it was implemented in this way. In terms of social validity, Wolf’s framework (17) will be utilised, focusing on key tenets of acceptability, feasibility, and utility (e.g., does the intervention meet schools’ perceived needs? How well received is the intervention among staff and pupils? Can the intervention be delivered successfully?). Relevant studies of school-based interventions (e.g.,) will be drawn up and existing rubrics from the implementation literature adapted (e.g.,) to inform data generation.

In terms of how MAPSS was implemented, the IPE will focus on the following dimensions: fidelity (e.g., to what extent teachers adhered to MAPSS guidance), dosage (e.g., how much of MAPSS pupils accessed), quality (e.g., how well MAPSS was delivered), participant responsiveness (e.g., the extent to which pupils engaged), reach (e.g., the rate and scope of participation), programme differentiation (e.g., to what extent MAPSS can be distinguished from other, existing mental health programmes), and adaptations (e.g., the nature and extent of changes made during implementation). A range of factors that may have affected implementation at the different domains/levels consistently will also be explored: preplanning and foundations (e.g. buy-in), implementation support system (e.g. ongoing external support), implementation environment (e.g. time constraints), implementer factors (e.g. experiences, skills and confidence in delivery), and programme characteristics (e.g. flexibility; (33–35)).

Longitudinal case study fieldwork visits will be conducted at T2 (after suicide prevention lesson) and T3 (after Reframe IT-UK). Semi-structured interviews will be used with school staff and intervention deliverers (N=3 per school x 1 visit = 12 interviews), and interviews (for Reframe IT-UK) and focus groups (for suicide prevention lesson) with pupils (N=1 per school x 1 visit = 4 focus groups; N=2 per school x 1 visit = 8 interviews), as well as observations and document analysis of intervention delivery.

Class teachers and members of the school senior leadership team (e.g., safeguarding leads) will be interviewed individually at each case study visit. Small groups (n=4-6) of pupils will participate in semi-structured focus groups regarding the suicide prevention lessons (to reduce power imbalances and ease nerves), and one-to-one interviews (with a teacher present if requested) will be conducted with pupils who have taken part in Reframe IT-UK (due to the sensitive and personal nature of intervention participation). Bespoke semi-structured interview schedules have been developed for each key stakeholder group. All interviews/focus groups will cover trial feasibility and acceptability, and factors affecting implementation; overarching this will be a social validity framework (17). However, each schedule will be tailored to the relevant time point and stakeholder group. Prompts and probes will be utilised where necessary to clarify unclear responses and elicit further detail. Interviews and focus groups will be conducted in private and quiet parts of the school, and fully informed consent/assent will be ensured.

Professionals who deliver the interventions in the case study schools will also be invited to be interviewed, to ascertain fidelity, quality, and dosage, and gain their perspectives on participant engagement and reach, as well as the feasibility of an efficacy trial. Interviews will be conducted at a time and place to suit them (face-to-face or online). Observations and document analysis will be arranged where possible with the intervention deliverers for additional context. All interviews/focus groups will be audio recorded and transcribed verbatim.

### Data Management

Source data for this trial will consist of paper copies of the consent form, data from online questionnaires (collected via QuestionPro), and audio recordings of interviews and focus groups.

When a participant consents to take part in the trial, they will be provided with a unique participant identification number which will be used to link survey data across timepoints. Personal data entered via the survey platform (QuestionPro) will be anonymised and stored on a password-protected database, housed on LJMU’s secure systems, and will only be accessible to members of the core research team. Consent forms and letters with personal identifiable data will be stored separately in a locked filing cabinet. Participant details will be anonymised in any publications that result from the trial.

Encrypted Dictaphones (or Microsoft Teams’ recording function if online) will be used to record interviews and focus groups. Audio files will be immediately transferred to LJMU’s secure servers after the interviews are complete and will subsequently be deleted from the Dictaphone or software platform. Once transcribed, audio files will also be deleted from LJMU’s systems. Transcripts will be anonymised, with any identifiable information removed, and pseudonyms used. Only the research team will have access to the transcripts. Direct access to data will be granted to authorised representatives from the Sponsor, host institution and the regulatory authorities to permit trial-related monitoring, audits and inspections - in line with participant consent.

This trial will be sponsored by LJMU, who are also the data custodian. All research data will be retained in a secure location during the conduct of the trial and for five years after the end of the trial, when all paper records will be destroyed by confidential means. An archiving plan will be developed for all trial materials in accordance with the LJMU archiving policy.

### Safety Considerations

The organisational structure of the trial is as follows. The trial steering committee (TSC) is responsible for investigator oversight of MAPSS and is comprised of the Co-Principal Investigators (Co-PIs; EA and PS), trial manager, clinical supervisor, and independent experts in the field. The TSC will meet on a six-monthly basis. The MAPSS Data Safety and Management Committee (DMEC) is the main vehicle for safety management, data monitoring, endpoint adjudication, and data management strategy. The committee is comprised of the Co-PIs and independent experts in the field. The composition of the group was designed to ensure transparency, so that no one set of competing interests could unduly influence other stakeholders, and is appropriate for this non-commercially funded or sponsored study. This committee has a dual safety role: it incorporates a risk-appropriate safety, endpoint adjudication, and data management strategy which is responsive to study issues as they eventuate. The committee will meet annually, and on an ad hoc basis when deemed necessary by the steering committee.

A comprehensive safety protocol has been developed, which will be activated if: 1) participants return a score of 21 or higher on the SIDAS at any timepoint (the established cut-off indicating a high level of suicidal ideation in the past month); 2) participants report current suicidal ideation at any timepoint; or 3) participants report suicide risk via the Reframe IT-UK platform. The school will be responsible for checking in with the participant and managing any risk. The response will depend on the level of risk, and ranges from providing the participant with contact information for helplines (in the case of low risk) to calling the participant’s emergency contact or emergency services (in the case of high-severe risk). Ultimately, all risk information will be communicated to the school, who will be responsible for ongoing management. Supervision will be provided to the researchers and school staff administering Reframe IT-UK from the study’s therapist. Adverse events (AEs) or serious adverse events (SAEs) that arise during the trial will be recorded in the study database.

### Assessment of Unanticipated Outcomes

An AE is the development of an untoward effect, undesirable clinical occurrence or medical condition, or the deterioration of a pre-existing medical condition following or during exposure to a study intervention, whether or not considered causally related to the study intervention. For the purposes of safety reporting, any research activity is considered to be part of the “study intervention”. An AE can therefore be any unfavourable and unintended clinical sign, symptom, observation, or disease temporally associated with the use of an intervention, whether or not related to the intervention. An SAE is any untoward medical occurrence that: results in death or is life-threatening (‘life-threatening’ in the definition of SAE refers to an event in which the participant was at risk of death at the time of the event, it does not refer to an event which hypothetically might have caused death if it were more severe); requires inpatient hospitalisation or prolongation of existing hospitalisation; results in a persistent or significant disability/incapacity; is an important medical event that although not immediately life-threatening or results in death/hospitalisation, based upon appropriate medical and scientific judgment, may jeopardise the participant and/or require intervention to prevent one of the outcomes listed above. Outpatient treatment in an emergency department is not in itself an SAE, although the reasons for it may be (e.g., suicide attempt). Hospital admissions and/or surgical procedures planned before or during a study are not considered SAEs, if the illness or disease existed (or the surgery was planned) before the participant was enrolled in the study, provided that it did not deteriorate in an unexpected way during the study.

All AEs and SAEs that arise during the trial will be recorded in the study database. Schools will be required to inform the research team of any AEs or SAEs that occur during the course of the trial. The causality of AEs and SAEs (i.e., their relationship to intervention treatment) will be assessed by a suitably qualified study team member. Any SAE will be reported to the Sponsor and to the relevant ethics committees within 24 hours of the research team becoming aware of its occurrence.

### Analytic Strategy

#### Quantitative Data

As this is a feasibility study no formal hypothesis testing will be undertaken. Data will be initially cleaned and checked for missing values; where possible missing values will be obtained from source or infilled using standard techniques, regression, or hot deck imputation, once all data have been collected. Demographics and other baseline variables will be reported using summary statistics, mean, medians, counts, or percentages depending on the nature (categorical or continuous) and distribution (parametric or non-parametric) of the data, along with corresponding measures of variability. Key outcomes from the study, for example recruitment and retention rates, will be reported using counts and percentages, along with 95% confidence intervals. If a 50% retention rate is assumed from an initial sample size of 810, it will be possible to estimate the true retention rate (95%) with an accuracy of +/-6%. Clinical outcomes, for example depression scores on a continuous scale, will be reported using means or medians depending on the distribution and corresponding confidence intervals. These clinical measures will also be reported at the three subsequent follow-up timepoints. Graphical methods will be used to identify trends across time. Differences between the intervention and the control groups in key outcome variables will also be calculated and reported graphically, along with 95% confidence intervals. All analysis will be carried out on an ITT basis, and some sensitivity analysis maybe undertaken including only per-protocol participants. Secondary outcomes will be reported and assessed in a similar manner to the primary outcomes using summary statistics and confidence intervals.

#### Qualitative Data

Qualitative data will be treated in two ways. First, detailed case profiles of each school will be produced that document their implementation, paying attention to how individual context and circumstances have influenced progress in each. Secondly, interview and focus group transcripts will be analysed via thematic analysis using the framework approach. (36). A hybrid approach will be taken, which will be informed by conceptual models of implementation in school settings (37) and our primary orienting concepts (social validity, acceptability, feasibility), while allowing for unanticipated themes specific to this project/context. Normalisation Process Theory (NPT; 35) will also be adopted as a broad framework through which to make sense of the qualitative data and draw conclusions relating to how readily MAPSS might be implemented amongst schools and embedded into school systems.

The qualitative framework analysis approach was developed to meet information needs and to provide outcomes or recommendations (39). It offers a highly visible and systematic approach to data analysis, showing very clearly how findings are derived from the data. This approach also facilitates analysis of specific concepts and issues that are particularly important to address, and so facilitates the use of NPT in interpreting the data. Analysis will follow the five suggested stages of framework analysis (Familiarisation with the data; Identifying a thematic framework; Indexing the data; Charting the data; Mapping and interpretation; 37).

NPT provides a framework for understanding the barriers and facilitating processes that underlie the implementation and integration of complex interventions into systems. The theory has been developed from qualitative research and identifies four key processes that underlie the adoption of new interventions (coherence of intervention; cognitive participation; collective action; reflexive monitoring). Previous research has shown that NPT can be applied effectively to qualitative data in healthcare contexts and, more recently, in school-based research (38,41,42). NPT will be drawn upon as a putative framework within the qualitative analysis, and an attempt will be made to map the links between qualitative themes arising from the data and the core processes outlined in NPT. This process will be aided through use of the NPT toolkit (http://www.normalizationprocess.org/) and application of the NPT statements generated by May et al. (38,42). In order to further promote integrity and rigor during the data analysis process, field notes will be written immediately after the interview and a reflective diary maintained.

#### Progression Criteria

A set of eight provisional stop/go progression criteria for the MAPSS programme have been established to determine whether a full RCT is warranted, and will be further developed in collaboration with the TSC (see Table 2). All progression criteria will need to be met for the MAPSS programme to be seen as acceptable and feasible, and to progress to a full efficacy trial.

**Table 2.**
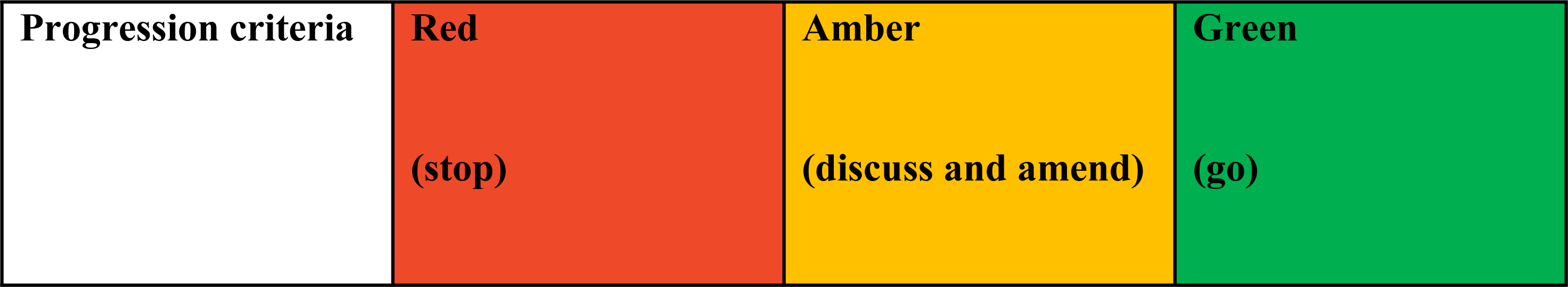
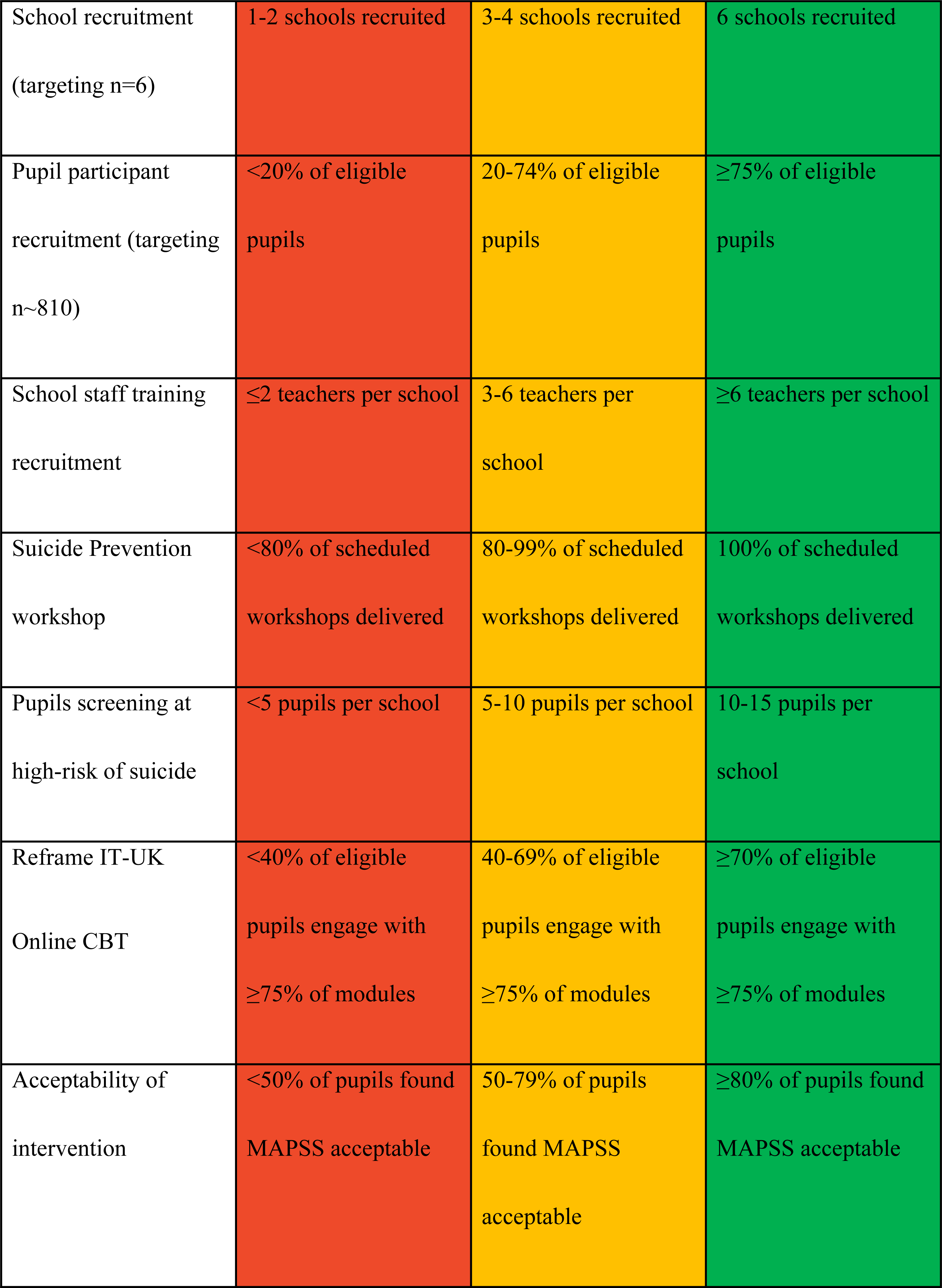
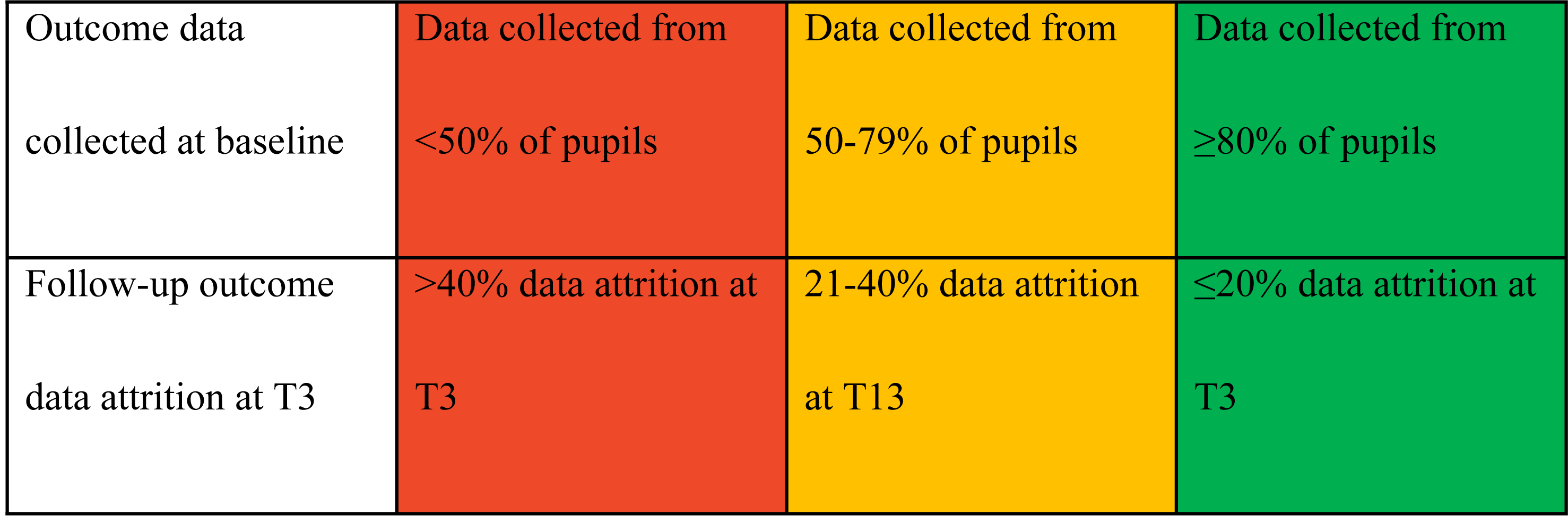
Trial progression criteria.

| Progression criteria | Red | Amber | Green |
| --- | --- | --- | --- |
|  | (stop) | (discuss and amend) | (go) |
| School recruitment<br>(targeting n=6) | 1-2 schools recruited | 3-4 schools recruited | 6 schools recruited |
| Pupil participant<br>recruitment (targeting<br>n~810) | <20% of eligible<br>pupils | 20-74% of eligible<br>pupils | ≥75% of eligible<br>pupils |
| School staff training<br>recruitment | ≤2 teachers per school | 3-6 teachers per<br>school | ≥6 teachers per school |
| Suicide Prevention<br>workshop | <80% of scheduled<br>workshops delivered | 80-99% of scheduled<br>workshops delivered | 100% of scheduled<br>workshops delivered |
| Pupils screening at<br>high-risk of suicide | <5 pupils per school | 5-10 pupils per school | 10-15 pupils per<br>school |
| Reframe IT-UK<br>Online CBT | <40% of eligible<br>pupils engage with<br>≥75% of modules | 40-69% of eligible<br>pupils engage with<br>≥75% of modules | ≥70% of eligible<br>pupils engage with<br>≥75% of modules |
| Acceptability of<br>intervention | <50% of pupils found<br>MAPSS acceptable | 50-79% of pupils<br>found MAPSS<br>acceptable | ≥80% of pupils found<br>MAPSS acceptable |
| Outcome data collected at baseline | Data collected from <50% of pupils | Data collected from 50-79% of pupils | Data collected from ≥80% of pupils |
| Follow-up outcome data attrition at T3 | >40% data attrition at T3 | 21-40% data attrition at T13 | ≤20% data attrition at T3 |

### Ethical Considerations

Ethical approval has been obtained from the university’s research ethics committee (REC; 23/PSY/003). The study will be undertaken in compliance with the research protocol. All participants will be given a participant information sheet and consent form prior to taking part. Personal data will be documented on password protected and encrypted servers housed at the research team’s institution. No identifiable patient data will be extracted.

As delivery of the intervention is being arranged by local Public Health bodies, consent will only be sought for completion of the measures. While opt-in gatekeeper consent will be sought from the participating schools, written opt-out consent will be sought from parents of CYP. Findings from both the scoping and pilot study consistently showed that opt-out consent is feasible and desirable for this project. Given the potentially sensitive nature of the measures, parents/carers will be informed of the project on two separate occasions (via the schools’ usual communication channels), to help ensure information is not missed.

Schools will also be asked to advise parents/carers of the date scheduled for survey completion, so they are aware. All parents will be provided with detailed information sheets (alternative format/easy-read will also be developed), outlining the importance of the study and any risk of harm (and procedures put in place to reduce this), and will be provided with detailed signposting. Parents will be able to view the items in the survey and attend an online information session about suicide prevention in CYP. Parents and carers who are Co-Investigators and Public and Patient Involvement (PPI) advisory members will be consulted to ensure 1) attendance at the parent information sessions, and 2) that parents/carers are effectively informed about any young people who may be at risk of suicide and equipped with appropriate resources and support.

Schools will be provided with a detailed support pack for completing the measures with pupils, including age-appropriate lesson plans, PowerPoint slides, and glossary. The slides inform the CYP of the nature of the study and their rights as a participant (including being able to withdraw) and can be delivered by the teacher supporting survey completion. CYP will then be able to indicate if they are happy to proceed by ticking a box at the beginning of the survey. This method has been used successfully in previous trials by the Principal Investigators, in the pilot study, and is recommended in good practice guidance.^38^ For pupils eligible for Reframe IT-UK, they will be provided with an information document/leaflet/video (co-developed with our young person’s advisory group), advising them about the content of Reframe IT-UK, and the voluntary nature of participation. The school’s guidance pack will also remind staff to ensure that pupils are provided information discretely and are made aware that they do not have to take part.

Fully informed written/verbal opt-in consent will be sought for participation in the qualitative strand of the IPE. Participants will be verbally reminded of their rights prior to the interviews/focus groups beginning. In case of distress to teachers during MAPSS, the school guidance packs will provide information on promoting staff wellbeing, including details of 24-hour helplines (one specifically for educators, and local NHS crisis lines). In case of distress to parents/carers during MAPSS, the parent/carer participant information sheets will provide details of charities e.g., Papyrus and NHS services including 24-hour crisis helplines.

#### Public and Patient Involvement (PPI)

To ensure the research meets the needs of, and is sensitive to, pupils and teachers in a school community, the proposed work has been developed as part of the Suicide and Self-Harm Research Group (SSHRG) at LJMU and with PPI co-applicants including parents and youth worker leads. PPI members have advised on overall study design, research questions, recruitment, and have helped write the plain English summary. The following approaches were taken towards involving the public in the development of this study:

- Liverpool NHS Clinical Commissioning Group (CCG) commissioned a qualitative scoping study to gain views from key stakeholders, including young people, parents of children with a history of suicidal behaviours, teachers, mental health professionals and General Practitioners. Data indicated strong support for MAPSS to bridge the current gap in clinical service provision and to support schools in managing/signposting the increasing number of young people communicating suicidal ideation. The scoping study informed the development of the trial design (e.g., trial recruitment procedures) and the adaptations required to MAPSS to ensure it meets the cultural needs of the UK population.
- Six parents were active members in the development of the feasibility study, with two as named co-applicants.
- A young person’s advisory group has been established via Merseyside Youth Association, who are co-designing information documents/leaflets/videos to empower young people, helping them to understand what both MAPSS and the CBT element will entail, and fully understand their rights throughout the process.
- The original Reframe IT intervention was developed with input from young people in Australia.

Moving forward in the feasibility trial, the research team will develop two advisory groups: one for adults (n=6) with lived experience of parenting/caring/working with young people with suicidal ideation (Public Advisory Group; PAG) and one for young people (n=6) with lived experience of suicidal ideation or with an interest in MAPSS (YPAG). The PPI Co-Is and PS will co-lead and co-ordinate the PAG. EA will co-lead and co-ordinate the YPAG, in collaboration with Merseyside Youth Association. The team will work with existing community links to ensure members from under-served communities (e.g., ethnic minority groups) are members. Involvement will be flexible and will use multiple methods to ensure members can engage according to their abilities and preferences. Members will have an induction and a discussion of working practices, delivered by the research team, who have extensive experience in involving CYP and adults as advisors in research. The team will invest time building trust with and develop safeguarding protocols for engagement. Members will advise the team on all elements of study conduct and dissemination, to ensure that findings are appropriately translated and are culturally sensitive and accessible. Funds (using NIHR rates) are included to ensure that all members may continue to support the research throughout the project.

Training will be available throughout the programme for PPI members and an induction to the project will be delivered. This will include an overview of the MAPSS programme and the methodology being used to evaluate the intervention, to enable the groups to be better informed about the project and their role in advising the research team. The ADAPT guidance (43) and Health Inequalities Assessment Toolkit (https://forequity.uk/hiat/) are being used to address the adaptation of this intervention in a different population and to review health inequalities and access for young people communicating/displaying suicidal behaviours.

For capturing, evaluating, and reporting the impact of PPI activities, the team will record minutes from all meetings. Focus groups will also be conducted with the PAG and YPAG to explore their experience of being part of the programme and use these findings to inform the effectiveness trial funding application.

#### Study Status and Timelines

Recruitment of schools and participants commenced 14^th^ February 2024 and will close when six schools have been recruited. The last participants will be followed-up at T4 in December 2025. Recruitment for the qualitative process evaluation will be ongoing through the course of the trial.

## Discussion

This study responds to the rising rates of suicide among young people in Northwest England but also, as noted above, a lack of high-quality evidence in youth suicide prevention research. In line with international best practice, this study will evaluate the feasibility and potential cost-effectiveness of a suicide prevention programme comprising universal, selective, and indicated components delivered in secondary school settings in the UK.

### Dissemination

Data arising from the trial is owned by the research team. On completion of the trial, the data will be analysed and tabulated, and a final trial report prepared, which will be available via the trial funder’s website.

Project findings will be shared through reports developed in close consultation with schools, NHS professionals, public health and third sector organisations, and those affected by suicide. Findings will be of interest to various stakeholder groups, and so bespoke reports will be developed for education, health and social care organisations, and researchers in the field. Outputs will include publications in high impact peer-reviewed journals; presentations/symposia at national and/or international conferences; summary briefs for different audiences; policy evidence briefings; a public-facing website, including short videos/animations, infographics; and blogs/vlogs to highlight the work.

A one-day national conference will be hosted, funded jointly (by organisations working on this project), focusing on dissemination and discussion of the project findings. Academics, researchers, schools, third sector organisations, social and clinical service staff, and the public will be invited, ensuring a range of voices and perspectives are present on the day. The one-day conference will be used as a platform to gain initial interest from NHS England Public Health Suicide Prevention Leads, through which future engagement can then be supported. Within the conference, individuals and carers affected by suicidal behaviours will be supported to participate, and their voices will be actively encouraged and listened to in considering the development and implementation of the subsequent trial.

Press releases at key project milestones will be disseminated via an ongoing social media campaign, designed to further disseminate project progress and findings. Summary and guidance documents will be created and made available to schools managing pupils with suicidal behaviours via the study website page. The next step of the research (efficacy RCT) will be supported by a pro-active engagement with schools across the region via NHS England Public Health Suicide Prevention Leads and the NIHR Clinical Research Network (CRN).

Study participants will be asked if they would like to be updated on forthcoming publications, and a note will be made of their responses.

### Study Amendments

Study procedures will not be changed without the mutual agreement of the Co-PIs, the trial funder, and the trial sponsor. If it is necessary for the study protocol to be amended, the amendment or a new version of the study protocol must be notified to or approved by the NIHR and REC before implementation, unless the safety of participants is at risk. The trial registry and the journal in which the protocol is published will be notified of any protocol updates.

## Data Availability

No datasets were generated or analysed during the current study. All relevant data from this study will be made available upon study completion.

## Acknowledgements

The present trial is funded by NIHR PHR programme. The previous scoping and pilot studies were funded by Liverpool NHS Clinical Commissioning Group and CHAMPS respectively. Papyrus Prevention of Young Suicide and Grassroots Suicide Prevention are delivering the MAPSS intervention in schools. We would like to thank all organisations for their contributions and ongoing collaborations. Finally, we would like to thank the public advisors who have contributed to the study design to date, including the ‘3 dads walking’ and Merseyside Youth Association.

## Metadata

### Funding

This research is funded by the National Institute for Health and Care Research (NIHR), Public Health Research Programme, UK (NIHR156862).

### Data Availability

Data will be available from the authors upon request at study completion.

